# *Chyawanprash* for the prevention of COVID-19 infection among healthcare workers: A Randomized Controlled Trial

**DOI:** 10.1101/2021.02.17.21251899

**Authors:** Arun Gupta, Amit Madan, Babita Yadav, Pallavi Mundada, Richa Singhal, Yogesh Kumar Pandey, Riju Agarwal, Arunabh Tripathi, Rakesh Rana, B. S. Sharma, BCS Rao, Bharti, Narayanam Srikanth, K. S. Dhiman

## Abstract

**Background:** Coronavirus disease 2019 (Covid-19) occurs after exposure to severe acute respiratory syndrome coronavirus 2 (SARS-CoV-2). For persons who are at high risk of exposure, the standard of care is personal protection from getting infected. Whether Ayurvedic *rasayana* drug like *Chyawanprash* can prevent symptomatic infection in frontline health care workers is unknown.

**Objective:** To evaluate the effect of the combination of *Chyawanprash* and Standard Preventive Regimen compared to the use of Standard Preventive Regimen alone on the proportion of RT-PCR confirmed COVID-19 infections among frontline healthcare workers (HCWs).

**Methods:** An open-label randomized controlled trial was conducted in the HCWs between 25 to 60 years age currently working in an environment with chance of direct exposure to COVID-19 cases. The interventions to be compared in this trial were Standard Preventive Regimen as per institutional guidelines and based on their roles (Group I) and Ayurvedic Intervention viz., *Chyawanprash* 12 g twice for 30 days from day of randomization plus Standard Preventive Regimen (Group II). The incidence of RT PCR confirmed COVID-19 cases in both groups, was the primary outcome measure. Evaluation of the safety of the study drug (by any statistically significant change in various biochemical and hematological parameters and occurrence of any adverse drug reactions); incidence of any other infective diseases (bacterial /viral/ fungal / etc.) like upper respiratory tract illness during the study period and any change in the immunoglobulins like IgG, IgM and IgE and inflammatory markers like TNF alpha, IL-6 and IL-10 were the secondary outcome measures.

**Results:** Out of 193 participants who completed the study, no participant in both groups was COVID-19 positive at the end of one month. In post intervention follow-up, 4 subjects in Group I and 2 subjects in Group II were COVID-19 positive. No adverse drug reaction or any serious adverse event was reported during the study. No clinically significant change in the safety parameters was observed before and after the study. Statistically significant rise in Serum IgG level was seen in Group II but other inflammatory and immune markers did not show statistically significant difference.

**Conclusion:** *Chyawanprash* was well tolerated by all the participants in the intervention group but to prove its adaptogenic effect and efficacy as an add-on to the standard care in preventing the occurrence of COVID-19, clinical trial for longer duration with larger sample size is needed.

**Trial registration:** Clinical Trials Registry of India *vide* CTRI/2020/05/025275 dated 20/05/2020

**Date of IEC approval:** 19.5.2020

## Introduction

### Background

COVID-19 is primarily transmitted from person-to-person through respiratory droplets. These droplets are released when someone with COVID-19 sneezes, coughs, or talks. Infectious droplets can land in the mouths or noses of people who are nearby or possibly be inhaled into the lungs. So, the best preventive measure is avoiding contact of the droplets until an effective vaccine is administered. Respiratory droplets can land on hands, objects or surfaces around the person when they cough or talk, and people can then become infected with COVID-19 from touching hands, objects or surfaces with droplets followed by touching their eyes, nose, or mouth as a habit. Such transmission of COVID-19 can also occur through droplets of those with mild symptoms or those who do not feel ill.^1^ Transmission due to short-range inhalation of aerosols is a possibility particularly in crowded medical wards and inadequately ventilated spaces. So, strict adherence to Infection Prevention Control (IPC) practices especially appropriate use of Personal protective equipment (PPE) is advised to protect the health care workers.^2^

Everyone is at risk of getting infected with the SARS CoV-2. In India, the first case of COVID-19 was documented on 31st January 2020.^3^ Till first week of January 2021, 10.4 million cases of COVID-19 were diagnosed in India, among them, 9.98 million have recovered and more than 150 thousand deaths have occurred due to COVID 19 in the country.^4^ Currently, care for patients with COVID-19 is primarily supportive. Care is given to patients to help relieve symptoms and manage respiratory and other organ failure. There are currently no specific antiviral treatments licensed for COVID-19, however many treatments are under investigation and there is limited evidence for the efficacy of these medicines, in the present situation.^5^

Till the arrival of the vaccine for prevention of COVID-19, numerous recommendations from the health authorities were issued as prophylactic measures. The Joint Monitoring Group and the National Task Force for COVID-19 management in India had recommended the prophylactic use of Hydroxychloroquine (HCQ) in asymptomatic health workers involved in containment and treatment of COVID-19 as well as in non-COVID hospitals/non-COVID areas of COVID hospitals/blocks; asymptomatic frontline workers, such as surveillance workers deployed in containment zones and paramilitary/police personnel involved in COVID-19 related activities and asymptomatic household contacts of laboratory confirmed cases with contraindications in retinopathy, Hypersensitivity to HCQ or 4-aminoquinoline compounds, G6PD deficiency, pre-existing cardiomyopathy and cardiac rhythm disorders, children under 15 years of age and in pregnancy and lactation. As per the revised advisory on the use of HCQ as prophylaxis for SARS-CoV-2 infection dated 23rd March, 2020, there is a rare possibility of cardiovascular side effects such as cardio-myopathy and rhythm (heart rate) disorders, transient visual disturbance including blurring of vision that needs the drug to be discontinued and so its intake is recommended to under strict medical supervision. Its use against COVID-19, even as a prophylaxis, needs continuous monitoring, especially in individuals with preexisting heart conditions.^6^ Chloroquine as prophylaxis is contraindicated in patients with severe renal or hepatic diseases.^7^ In context with controlling the spread of disease, state agencies also undertook population-wide distribution of yet unproven homeopathic and Ayurvedic medicines and herbal tea mixes (ukalo), claiming they boost immunity and prevent quarantined individuals from getting infected.^8^ Practitioners also prescribed various other medications, including the anti-parasitic drug ivermectin.^9^ Ivermectin is currently not recommended in the national guidelines but can be used in patients in whom HCQ is contraindicated as per the expert opinion published on website of Ministry of Health and Family Welfare (MoHFW), Government of India.^10^

Although there is high risk of exposure during health care in the isolation wards, in the background of community transmission, there is always a high risk of transmission to the healthcare workers (HCWs) from pre-symptomatic and asymptomatic patients reporting with non-COVID illness also in non-COVID-19 hospitals.^11^ While the COVID-19 care hospitals are equipped with specialized safety measures and PPEs, doctors who treat patients before the hospitalisation stage (general practitioners or family physicians) are not protected similarly. N95 masks and gloves are the protective equipments recommended by the Government for the outpatient doctors. But the HCWs who deal with patients prior to confirmed diagnoses, using mere masks and gloves may not be protected sufficiently. Several patients hide the history of their exposure and triaging patients into fever and non-fever categories is also not done in small and congested clinics eventually, the healthcare workers pose more threat of getting infected in such a scenario. The recent news report reveals that more than 87,000 healthcare workers have been infected with COVID-19 with 9% positivity rate, with just six states viz., Maharashtra, Karnataka, Tamil Nadu, Delhi, West Bengal, and Gujarat accounting for three-fourths of the case burden and over 86% of the 573 deaths among the healthcare workforce. Possible factors responsible for high infections include lax infection control in healthcare facilities and the lack of stringent containment measures in areas where healthcare workers reside.^12^

Therefore, although the standard of care to prevent exposure to the virus containing droplets is of utmost importance to avoid spread of COVID-19, there is a need to gather evidence for effectiveness of certain interventions that could be safely co-administered to enhance immunity and build strength in the host to prevent the infection especially in high risk individuals. So this study was conducted to evaluate the efficacy of an Ayurvedic formulation which is categorized as a *Rasayana* medicine, as an add on to the standard preventive measures, in preventing COVID-19 infection in health care workers functioning at a COVID-19 care hospital in Delhi.

### Objective of the study

The present study was conducted to assess the impact of *Chyawanprash* on incidence of SARS-CoV-2 (COVID 19) infection among health care personnel exposed to COVID-19 cases.

## Methods

### Study Design

A randomized controlled trial was conducted between May 2020 to September 2020 among 199 HCWs functional at the COVID-19 Isolation Ward in Choudhary Brahmaprakash Ayurveda Charak Sansthan’s Hospital, Khera Dabur, New Delhi, India. The study was conducted in compliance with applicable ethical guidelines. Ethics committee approvals were obtained before study initiation. The Study was registered with the Clinical Trail Registry of India *vide* CTRI/2020/05/025275 dated 20/05/2020. The CONSORT statement guidelines have been followed in reporting the outcomes of the study.

### Study Participants

All the Health care workers of either sex functional at the said study setting during the study period were screened for inclusion and exclusion criteria after getting informed written consent.

### Inclusion Criteria

All HCWs between 25 to 60 years currently working in an environment with direct exposure to patients with confirmed COVID-19 infection were eligible to participate in the trial.

### Exclusion Criteria

HCWs who declined consent, who had a confirmed COVID-19 infection, those who were already taking chloroquine/HCQ for any indication or any other prophylactic drug, pregnant or breast-feeding women, having known co-morbidity or immune-compromised state, having known allergy for the study drug were excluded.

### Randomization

Eligible participants were allocated 1:1 in either group randomly by using a computer generated list of random numbers. This list was generated at the headquarters of Central Council for Research in Ayurvedic Sciences, New Delhi. This was an open label study. Assigned treatment was known to the research team and participant. Bias was supposed to be mitigated through an objective end point (laboratory confirmed COVID-19 infection).

### Study Intervention

The interventions to be compared in this trial were Standard Preventive Regimen (Group I) and Ayurvedic Intervention viz., *Chyawanprash* 12 g twice plus Standard Preventive Regimen (Group II). *Chyawanprash* is a classical Ayurvedic formulation. The Standard Preventive Regimen included standard precautions like Hand hygiene, Personal protective equipment (as per institutional guidelines and based on their roles), Respiratory hygiene and cough etiquette. Participants were discouraged from taking any other home remedies or other preventive measures in both groups (such as gargling with hot water etc.) throughout the study period. In the current study *Chyawanprash* manufactured by Dabur India Limited was used.

### Study Procedure

Participants were assessed clinically on Baseline, 7^th^ day, 15^th^ and 30^th^ day for protocol compliance and recording any adverse events if happened. As the duration of intervention was 30 days from the day of randomization, the lab investigations were done at the baseline and on 30^th^ day.

### Outcomes

The primary outcome was assessment of prophylactic efficacy of Chyawanprash as add on intervention to the standard preventive measures adopted by HCWs in a COVID care hospital. So the primary outcome measure was incidence of COVID-19 cases confirmed by Reverse Transcriptase Polymerase Chain Reaction (RT-PCR) test, in both the groups.

The secondary outcomes were evaluation of safety of the study drug by comparing the biochemical and hematological parameters before and after the study and through occurrence of any adverse drug reactions, assessment of efficacy of Chyawanprash in preventing other infective diseases (bacterial /viral/ fungal / etc.) like Upper respiratory tract illness through incidence of group of symptoms like Fever/fatigue/cough/anorexia/malaise/muscle pain/sore throat/dyspnoea/nasal congestion/headache and evaluation of effect of Chyawanprash on immunoglobulins and inflammatory markers through comparing the levels of IgG, IgM, IgE, high sensitivity C-Reactive Protein (hsCRP), Tumor Necrosing Factor alpha (TNF alpha) and Interleukins viz., IL-6 and IL-10. The immunoglobulins and inflammatory markers were assessed in every fifth subject in each group before and after the study.

### Statistical Analysis

#### Sample size calculation

On the basis of assuming incidence of COVID – 19 in only 40% patients in Group I (Standard of care) as compared to 20% in Group II (*Chyawanprash* as add on to the standard of care) with 95% Confidence Level (α = 0.05), 80% power and expecting a dropout rate of 20%, the number of patients to be enrolled in the study was calculated as approximately 100 in each group. Therefore, total sample size was 200 health care workers.

Data was collected in pre-designed Case Report Forms (CRFs) and entered in an electronic format prepared in ms-excel. Qualitative variables are described in Number (%) while quantitative variables are described in mean (SD) or Median (Q1, Q3). Qualitative variables in the study have been compared using chi-square test. The quantitative variables are compared by repeated measure ANOVA/Friedman test and paired t test/Wilicoxon sign test in within group. In between group comparison t test/Mann Whitney test is used. The level of significance is taken at 5%.

## Results

Out of total 204 HCWs screened, 199 were enrolled in the study. 95 participants in the control group (Group I) and 98 in the intervention group (Group II) completed the study (Fig. No.1). In Group I, 04 participants were drop outs as they didn’t want to continue participation and 02 participants in Group II were lost to follow up as they were not able to visit for follow-up.

**Fig. No.1.**
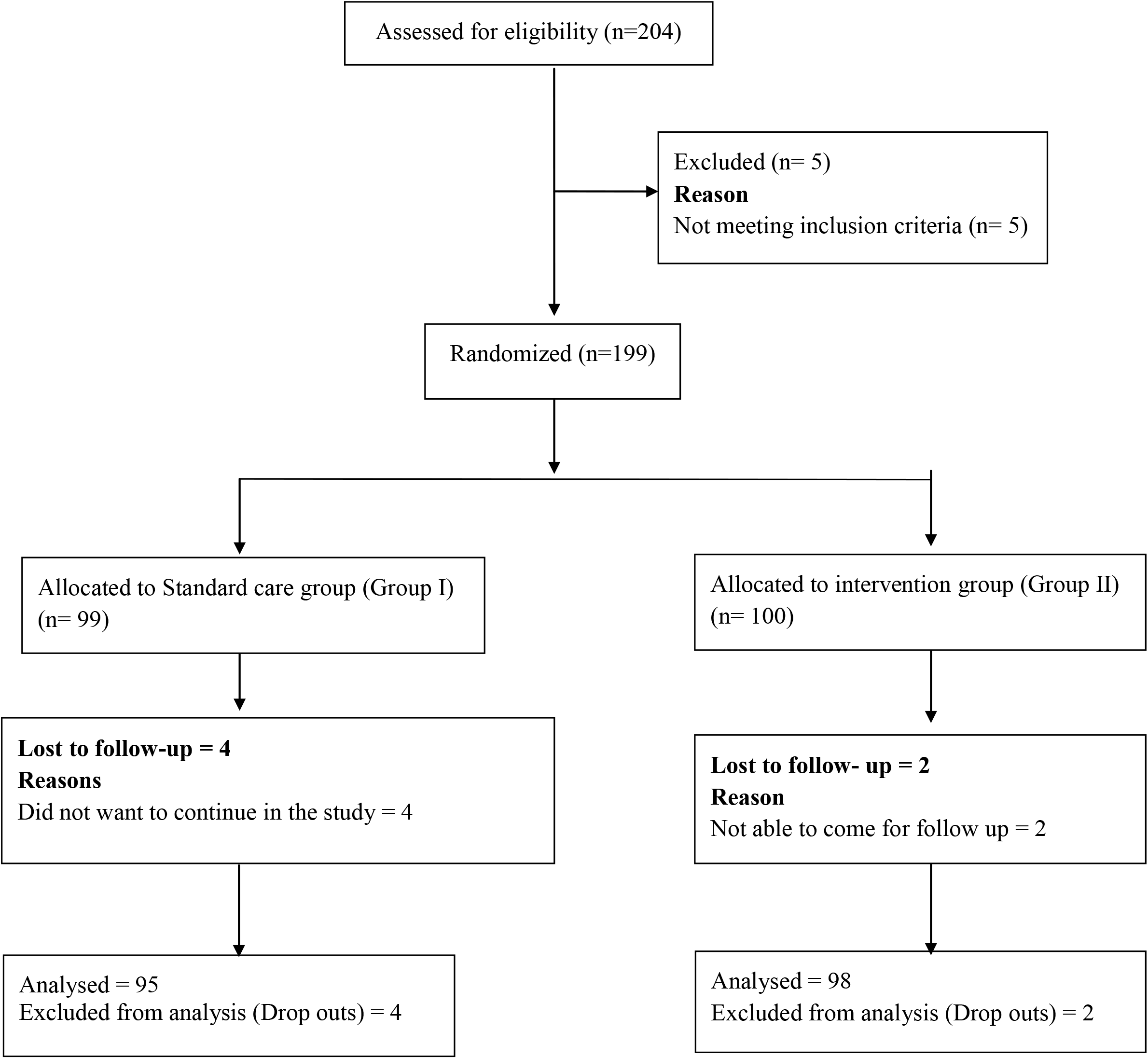
Flow diagram describing screening and randomization

### Demographic and clinical characteristics

Baseline characteristics of the participants in both the groups were similar [Table 1]. In the standard care group that is Group I, 50.5% participants were female, mean age was 33.35 years and mean Body Mass Index (BMI) was 24.44 kg/square meter. Whereas in Group II, in which participants were given *Chyawanprash* along with standard protective measures, 59.2% were male, mean age was 32.12 years and mean BMI was 23.56 kg per square meter. History of allergy to some material was reported by 14.7% participants in Group I and 17.3% in Group II. Majority of the participants in both groups had no addiction history. None of the participants had history of COVID-19 in the family prior to enrollment in both groups. In group I almost 30% participants had moderate to too much stress whereas in Group II 17% participants reported having moderate to too much stress. However this difference between both groups is not significant statistically. The participants with different work profiles were enrolled and equally distributed in two groups. [Table 1]

**Table No. 1.** Baseline characteristics of the participants in both the groups.

| Assessment Parameter | Group I (Control Group)<br>(n=95) | Group II (Intervention Group)<br>(n=98) | *p-value* |
| --- | --- | --- | --- |
| **Gender** |  |  |  |
| Male | 47 (49.5) | 58 (59.2) | 0.176 |
| Female | 48 (50.5) | 40 (40.8) |  |
| **Age (years)** | 33.357 (8.567) | 32.122 (7.390) | 0.284 |
| **BMI (kg/square meter)** | 24.440 (3.817) | 23.564 (3.748) | 0.110 |
| **Marital status** |  |  |  |
| Married | 64 (67.4) | 63 (64.3) | 0.708 |
| Unmarried | 29 (30.5) | 34 (34.7) |  |
| Widower | 2 (2.1) | 1 (1.0) |  |
| **Addictions** |  |  |  |
| Smoking | 6 (6.3) | 3 (3.1) | 0.341 |
| Alcohol | 8 (8.4) | 13 (13.3) |  |
| None | 81 (85.3) | 82 (83.7) |  |
| **Allergy to some material (Present)** | 14 (14.7) | 17 (17.3) | 0.622 |
| **Any emotional stresses** |  |  |  |
| Average | 65 (69.1) | 81 (82.7) | 0.080 |
| Moderate | 26 (27.7) | 16 (16.3) |  |
| Too much | 3 (3.2) | 1 (1.0) |  |
| **Nutrition** |  |  |  |
| Moderately Nourished | 46 (48.4) | 56 (57.1) | 0.157 |
| Malnourished | 5 (5.3) | 1 (1.0) |  |
| Well Nourished | 44 (46.3) | 41 (41.8) |  |
| **Work Profile** |  |  |  |
| Doctors | 29 (30.53) | 31 (31.63) | 0.745 |
| Nurses | 13 (13.68 ) | 17 (17.35) |  |
| Other paramedical staff | 39 (41.05 ) | 40 (40.82) |  |
| Housekeeping staff | 14 (14.74) | 10 (10.20) |  |
| All the values have been represented as n (%), except age and BMI which have been shown as Mean (SD)<br>Compared using Pearson Chi-square test<br>* *p-value* of <0.05 has been considered as significant |  |  |  |

### Assessment of Efficacy

None of the participant in either group was found COVID-19 positive at the end of study period. As the study revealed that no participant in any group got infected from COVID-19, we telephonically followed the participants for next four months after completion of study period for knowing the protection rate in both the groups. This follow-up data regarding incidence of COVID-19 among the study participants revealed that, 04 participants in Group I showed symptoms like fever, cough, anosmia and loss of taste and tested COVID-19 positive as confirmed by RT-PCR, out of them 01 had to be hospitalized, whereas only 02 participants in Group II tested COVID-19 positive and both were asymptomatic. All the 6 participants were diagnosed as COVID-19 positive within 2 months after completion of study period.

There was no incidence of any other infective diseases (bacterial /viral/ fungal / etc.) like upper respiratory tract illness during the study period in both the groups.

Assessment of inflammatory and immune markers in 18 participants in each group showed that median IgG levels increased significantly from 1337.5 (1185, 1610.4) at baseline to 1361 (1126, 1675) on 30^th^ day in Group II (*p*=0.016). Median IgE levels decreased significantly in Group I from 173 (90.5, 420) at baseline to 159 (79, 424.5) on 30^th^ day (*p*=0.019), however the values were within normal limits. All the other immune and inflammatory markers like IgM, hsCRP, IL6, IL10 and TNF alpha did not show statistically significant changes before and after study period in any group and all the values were within normal limits. [Table 2]

**Table No. 2.** Assessment of Immune and Inflammatory markers.

| Laboratory Parameters | Group I (n=18) | Group II (n=18) | *p-value*<sup>\$</sup> |
| --- | --- | --- | --- |
| **hsCRP (mg/litre)** |  |  |  |
| Baseline | 1.5 (0.325, 2.680) | 0.73 (0.31, 1.335) | 0.066 |
| 30<sup>th</sup> Day | 1.365 (0.947, 3.31) | 1.075 (0.772, 1.86) | 0.268 |
| *p-value*<sup>#</sup> | 0.463 | 0.184 |  |
| **IL-6 ( <141.9 pg/ml)** |  |  |  |
| Baseline | 52.05 (13.987, 201.02) | 26.3 (7.68, 194.05) | 0.624 |
| 30<sup>th</sup> Day | 46.45 (6, 199.25) | 18.9 (1.425, 124.12) | 0.496 |
| *p-value*<sup>#</sup> | 0.535 | 0.795 |  |
| **IL-10 (<76.3 pg/ml)** |  |  |  |
| Baseline | 16.900 (5.512, 46.025) | 16.8 (4.842, 30.475) | 1.000 |
| 30<sup>th</sup> Day | 19.75 (14.75, 57.7) | 21.2 (11.4, 32.45) | 0.962 |
| *p-value*<sup>#</sup> | 0.698 | 0.446 |  |
| **TNF alpha (<108.5 pg/ml)** |  |  |  |
| Baseline | 26.4 (8.85, 94.05) | 28.3 (16.575, 168.2) | 0.548 |
| 30<sup>th</sup> Day | 14.9 (9.2, 46.15) | 14.1 (7.075, 46.35) | 0.874 |
| *p-value*<sup>#</sup> | 0.182 | 0.215 |  |
| **IgG (700-1600 mg/dl)** |  |  |  |
| Baseline | 1447.65 (1099.2, 1585.5) | 1337.5 (1185, 1610.4) | 0.887 |
| 30<sup>th</sup> Day | 1365.5 (1164.2, 1680.5) | 1361 (1126, 1675) | 0.924 |
| *p-value*<sup>#</sup> | 0.193 | 0.016* |  |
| **IgM (40-230 mg/dl)** |  |  |  |
| Baseline | 88.8 (61.675, 133.98) | 119.95 (78.15, 189.75) | 0.137 |
| 30<sup>th</sup> Day | 89.5 (65.425, 162) | 143 (87.25, 234.25) | 0.103 |
| *p-value*<sup>#</sup> | 0.093 | 0.381 |  |
| **IgE (0.4-378 IU/ml)** |  |  |  |
| Baseline | 173 (90.5, 420) | 160.55 (49, 445.5) | 0.812 |
| 30<sup>th</sup> Day | 159 (79, 424.5) | 183.5 (55.75, 355.25) | 0.837 |
| *p-value*<sup>#</sup> | 0.019* | 0.868 |  |
| Values have been represented as Median (Q1, Q3)<br># Compared using Wilcoxon sign rank test<br>\$Compared using Mann-Whitney test<br>* *p-value* of <0.05 has been considered as significant | | | |

### Assessment of Safety

None of the participant in either group was found COVID-19 positive at the end of study period. No adverse event was recorded during the study. The vital parameters of all the participants were within normal limits in both groups throughout the study. Statistically significant decrease in mean systolic blood pressure was observed in Group II at the end of study period. However, the change is not biologically significant [Table 3]. All the hematological parameters in both groups were within normal limits before and after the study [Table 4]. Mean blood urea, serum creatinine and SGOT levels were decreased in both the groups on 30^th^ day, while total protein, serum albumin and SGPT levels decreased and serum globulin increased in group II on 30^th^ day. However, all these values were within normal limits at baseline as well as on 30^th^ day [Table 5].

**Table No. 3.** Assessment of vital parameters.

| Assessment parameters | Baseline | 7<sup>th</sup> Day | 15<sup>th</sup> Day | 30<sup>th</sup> day | *p-value*# |
| --- | --- | --- | --- | --- | --- |
| Respiratory Rate (per minute) |  |  |  |  |  |
| Group I (N=95) | 17.357 (1.999) | 16.978 (1.862) | 17.378 (1.805) | 17.052 (1.847) | 0.065 |
| Group II (N=98) | 17.061 (2.055) | 16.663 (1.931) | 16.693 (2.737) | 16.734 (2.103) | 0.369 |
| *p-value*\$ | 0.311 | 0.249 | 0.042* | 0.266 | |
| Pulse Rate (per minute) |  |  |  |  |  |
| Group I (N=95) | 76.642 (5.040) | 76.526 (4.614) | 75.642 (8.805) | 76.452 (4.533) | 0.539 |
| Group II (N=98) | 75.836 (3.942) | 75.581 (4.490) | 74.969 (9.179) | 75.806 (4.085) | 0.631 |
| *p-value*\$ | 0.217 | 0.151 | 0.604 | 0.299 | |
| Systolic Blood Pressure (mm of Hg) |  |  |  |  |  |
| Group I (N=95) | 119.13 (8.351) | 117.37 (13.401) | 118.46 (8.724) | 116.4 (8.488) | 0.136 |
| Group II (N=98) | 120.18 (6.802) | 118.21 (6.642) | 118.88 (7.128) | 117.04 (6.881) | 0.004* |
| *p-value*\$ | 0.335 | 0.577 | 0.718 | 0.565 | |
| Diastolic Blood Pressure (mm of Hg) |  |  |  |  |  |
| Group I (N=95) | 78.736 (6.451) | 77.768 (6.383) | 77.442 (6.750) | 78.021 (6.931) | 0.355 |
| Group II (N=98) | 78.959 (5.289) | 78.734 (6.087) | 79.051 (6.717) | 78.612 (6.033) | 0.943 |
| *p-value*\$ | 0.793 | 0.283 | 0.099 | 0.528 | |
| Values have been represented as Mean (SD) |  |  |  |  |  |
| #Compared using Repeated Measure Anova / Friedman test |  |  |  |  |  |
| \$Compared using independent sample t-test | | | | | |
| * *p-value* of <0.05 has been considered as significant |  |  |  |  |  |

**Table No. 4.** Assessment of Haematalogical parameters.

| Laboratory Parameters | Group I (n=95) | Group II (n=98) | *p-value*<sup>\$</sup> |
| --- | --- | --- | --- |
| **Haemoglobin (13.0 - 17.0 g/dl)** |  |  |  |
| Baseline | 13.338 (1.921) | 13.674 (1.934) | 0.228 |
| 30<sup>th</sup> Day | 13.360 (2.010) | 13.906 (1.993) | 0.060 |
| *p-value*<sup>#</sup> | 0.916 | 0.249 |  |
| **Total Leucocyte Count (4.0 - 10.0 10<sup>3</sup>per microlitre)** |  |  |  |
| Baseline | 7.193 (1.735) | 7.144 (1.697) | 0.844 |
| 30<sup>th</sup> Day | 7.180 (1.618) | 7.385 (2.206) | 0.464 |
| *p-value*<sup>#</sup> | 0.946 | 0.293 |  |
| **Neutrophils (40-80 %)** |  |  |  |
| Baseline | 53.547 (8.549) | 52.367 (7.593) | 0.312 |
| 30<sup>th</sup> Day | 55.452 (8.200) | 54.091 (8.146) | 0.249 |
| *p-value*<sup>#</sup> | 0.056 | 0.057 |  |
| **Eosinophils (1 – 6 %)** |  |  |  |
| Baseline | 2.852 (1.406) | 2.9184 (1.382) | 0.744 |
| 30<sup>th</sup> Day | 2.757 (1.513) | 3.020 (1.392) | 0.211 |
| *p-value*<sup>#</sup> | 0.631 | 0.608 |  |
| **Lymphocytes (20 – 40 %)** |  |  |  |
| Baseline | 36.515 (7.532) | 37.959 (6.954) | 0.168 |
| 30<sup>th</sup> Day | 36.178 (8.467) | 37.102 (7.420) | 0.421 |
| *p-value*<sup>#</sup> | 0.701 | 0.287 |  |
| **Absolute Lymphocyte Count (710 - 4530/mm<sup>3</sup>)** |  |  |  |
| Baseline | 2637.7 (720.573) | 2689.8 (663.547) | 0.602 |
| 30<sup>th</sup> Day | 2570.6 (711.571) | 2666.9 (714.131) | 0.349 |
| *p-value*<sup>#</sup> | 0.446 | 0.757 |  |
| **ESR (2 – 25 mm/hour)** |  |  |  |
| Baseline | 12.0 (8.0, 30.0) | 10.0 (8.0, 20.0) | 0.128 |
| 30<sup>th</sup> Day | 12.0 (8.0, 18.0) | 12.0 (8.0, 18.0) | 0.840 |
| *p-value*<sup>#</sup> | 0.002* | 0.525 |  |
| Values have been represented as Mean (SD) / Median (Q1, Q3)<br># Compared using paired sample t-test / Wilcoxon sign rank test<br>\$Compared using independent sample t-test / Mann Whitney Test<br>* *p-value* of <0.05 has been considered as significant | | | |

**Table No. 5.** Assessment of Biochemical parameters.

|  | **Group I (n=95)** | **Group II (n=98)** | **p-value** |
| --- | --- | --- | --- |
| **Fasting Blood Sugar (70 - 110 mg/dl)** |  |  |  |
| Baseline | 92.694 (14.941) | 89.724 (11.476) | 0.122 |
| 30<sup>th</sup> Day | 93.589 (19.461) | 89.102 (14.866) | 0.073 |
| **p-value<sup>#</sup>** | 0.676 | 0.736 |  |
| **Blood Urea (19 – 42 mg/dl)** |  |  |  |
| Baseline | 22.968 (5.667) | 22.234 (4.781) | 0.332 |
| 30<sup>th</sup> Day | 21.183 (5.078) | 21.071 (5.488) | 0.884 |
| **p-value<sup>#</sup>** | 0.005* | 0.033* |  |
| **Serum Uric Acid (3.1 - 8.2 mg/dl)** |  |  |  |
| Baseline | 4.83 (1.415) | 4.63 (1.358) | 0.331 |
| 30<sup>th</sup> Day | 4.85 (1.474) | 4.66 (1.197) | 0.267 |
| **p-value<sup>#</sup>** | 0.880 | 0.994 |  |
| **Serum Creatinine (0.4-1.20 mg/dl)** |  |  |  |
| Baseline | 0.825 (0.168) | 0.795 (0.183) | 0.236 |
| 30<sup>th</sup> Day | 0.773 (0.195) | 0.744 (0.211) | 0.322 |
| **p-value<sup>#</sup>** | 0.028* | 0.049* |  |
| **Total protein (6.0 - 8.0 gm/dl)** |  |  |  |
| Baseline | 7.357 (0.821) | 7.281 (1.288) | 0.626 |
| 30<sup>th</sup> Day | 7.413 (1.043) | 7.592 (0.99) | 0.223 |
| **p-value<sup>#</sup>** | 0.673 | 0.049* |  |
| **Serum Albumin (4.0 - 5.3 gm/dl)** |  |  |  |
| Baseline | 4.649 (0.327) | 4.720 (0.319) | 0.13 |
| 30<sup>th</sup> Day | 4.707 (0.434) | 4.638 (0.35) | 0.228 |
| **p-value<sup>#</sup>** | 0.286 | 0.040* |  |
| **Serum Globulin (2.5 - 3.8 gm/dl)** |  |  |  |
| Baseline | 3.041 (1.198) | 2.455 (0.088) | 0.086 |
| 30<sup>th</sup> Day | 2.741 (0.956) | 3.275 (0.343) | 0.15 |
| **p-value<sup>#</sup>** | 0.402 | 0.016* |  |
| **SGOT/AST (10 – 40 IU/L)** |  |  |  |
| Baseline | 31.0 (25.0, 40.0) | 32.0 (24.0, 43.0) | 0.985 |
| 30<sup>th</sup> Day | 29.0 (23.0, 40.0) | 29.5 (21.0, 38.0) | 0.755 |
| **p-value<sup>#</sup>** | 0.031* | 0.033* |  |
| **SGPT/ALT (10 – 40 IU/L)** |  |  |  |
| Baseline | 32.0 (23.0, 56.0) | 34.0 (20.7, 59.5) | 0.818 |
| 30<sup>th</sup> Day | 33.0 (22.0, 56.0) | 29.0 (21.0, 51.2) | 0.334 |
| **p-value<sup>#</sup>** | 0.902 | 0.013* |  |
| **Alkaline phosphatase (53.0-128.0 IU/L)** |  |  |  |
| Baseline | 112.11 (31.704) | 111.23 (34.715) | 0.762 |
| 30<sup>th</sup> Day | 116.08 (44.089) | 117.84 (45.400) | 0.787 |
| **p-value<sup>#</sup>** | 0.331 | 0.133 |  |
| **Serum Bilirubin Conjugated (0.1 - 0.3 mg/dl)** |  |  |  |
| Baseline | 0.226 (0.102) | 0.210 (0.098) | 0.279 |
| 30<sup>th</sup> Day | 0.281 (0.640) | 0.245 (0.348) | 0.630 |
| **p-value<sup>#</sup>** | 0.331 | 0.283 |  |
| **Serum Bilirubin unconjugated (0.2-1.20 mg/dl)** |  |  |  |
| Baseline | 0.455 (0.328) | 0.493 (0.4) | 0.476 |
| 30<sup>th</sup> Day | 0.430 (0.248) | 0.471 (0.3) | 0.309 |
| **p-value<sup>#</sup>** | 0.312 | 0.587 |  |
| Values have been represented as Mean (SD) / Median (Q1, Q3) |  |  |  |
| # Compared using paired sample t-test / Wilcoxon sign rank test |  |  |  |
| \$Compared using independent sample t-test / Mann Whitney Test | | | |
| *p-value of <0.05 has been considered as significant |  |  |  |

## Discussion

The study was based on the hypothesis that *Chyawanprash* could potentially be used as a safe prophylactic intervention along with standard precautionary measures, to prevent symptomatic infection in population having high risk of exposure to SARS CoV-2 virus. But the protective effect of *Chyawanprash* as an add on to the standard of care among HCWs functional at Isolation ward in a COVID-19 Care hospital, could not be proved primarily in the present study as there was no incidence of COVID-19 case in any group at the end of study period i.e. on 30^th^ day. However, in the long term follow-up after completion of the intervention period, the participants in the study group were found to be more protected from severe infection of COVID-19 as only 2 participants were RT-PCR positive after 2 months of completion of study period and both were asymptomatic. Whereas in the control group 4 participants were RT-PCR positive in same duration, and one out of them had to be hospitalized.

In the current study, the immunomodulatory and adaptogenic effect of *Chyawanprash* was proposed to be assessed through comparing the change in inflammatory and immune markers in every fifth participant in both groups before and after the study. IgG levels increased significantly among the participants taking *Chyawanprash* however, clinically the participants were normal. Serum IgG is an indicator of the humoral immune response. Its level represents the natural antibodies against antigens commonly encountered by the individual. IgG is the predominant antibody in the serum and it carries the major burden of neutralizing bacterial toxins and binding to microorganisms to enhance their phagocytosis. The values of IgM, hsCRP, IL6, IL10 and TNF alpha did not change significantly in any group in this study. In previous clinical trials conducted to explore the immune-modulatory, adaptogenic and cytoprotective effect of *Chyawanprash*, it is observed that, significant change in the immunity markers happens when it is given in higher dose and for longer duration. [Table No. 6] The classical dose for *avaleha* (linctus) preparation is almost 48 gram (as mentioned, 1 *pala* in Sharangdhar Samhita) but, *Rasayana* medicines are to be administered as per the digestive capacity and strength of the recipient. So in the present study, the standard dose of *Chyawanprash* as mentioned in the Ayurvedic Pharmacopoeia of India (Vol. 1, Part II) ^19^ was administered that is 12 gm twice daily and the duration of the intervention in the study only for 30 days. So this study might not have shown results to prove its immunomodulatory effect as observed in previous clinical trials.

**Table No. 6.** Review of previous studies exploring adaptogenic effect of *Chyawanprash*.

| Authors, Journal and Year of Publication | Population studied and total sample size | Dose of *Chyawanprash* administered | Duration | Outcomes |
| --- | --- | --- | --- | --- |
| <sup>13</sup> A. N. Uma & Dhananjay S. Kotasthane<br>Journal of Oral Oncology/ 2014 | 21 betel quid chewing oral precancerous lesions patients | 20 gm twice a day | 03 months | Comparison of the frequency of satellite association in the lymphocyte cultures before and after Cp feed after quitting betel quid chewing<br>58.73(14.90)- 25.20(6.83) ($p$ 0.0001)<br>After cessation of Betel quid chewing, Cp can further minimize the genotoxic effect caused by mutagenic agents present in betel quid. |
| <sup>14</sup> J.S. Yadav, S. Thakur & P. Chadha<br>International Journal of Human Genetics/ 2003 | 25 Male bidi smokers | 20 gm twice a day | 02 months | CP along with continued smoking habit, showed that Mitotic Index was significantly lower in Cp-fed bidi smokers (5.15) as compared with bidi smokers (6.36) ($P$ <0.01)<br>The frequency of Chromosomal Aberrations in Cp-fed bidi smokers (1.00) was significantly less than the same in bidi smokers (3.52) ($P$ < .001)<br>Frequency of sister chromatid exchanges in bidi smokers before (8.07) and after the administration of Cp (6.50) was significantly decreased ($P$<0.005)<br>Frequency of Satellite Association was decreased in the Cp-fed bidi smokers (7.12), almost to the level of control individuals (5.36) |
| <sup>15</sup> N. Gupta and S.C. Mahapatra<br>Proc. 2<sup>nd</sup> International Symposium on Medicinal and Nutraceutical Plants Acta Hort. 972, ISHS/ 2013 | 12 healthy adult males | 15 gm per day | 08 weeks | Significant increase in the erythrocyte superoxide dismutase (an antioxidant enzyme) activity from $1.41 \pm 0.16 \times 10^3$ to $1.77 \pm 0.29 \times 10^3$ U/gmHb ($p$<0.05)<br>serum IgG levels decreased from 16.4 to 11.4 g/L ($p$<0.05) |
| <sup>16</sup> P. K. Debnath et al. Journal of Ayurveda & Integrative Medicine/ 2012 | 99 newly diagnosed PTB patients from both the sexes aged between 10 and 65 years. | 10 gm thrice daily | 28 days | Normalization of erythrocyte sedimentation rate, appreciable change in IgA and IgM patterns and reduced bacterial load significantly earlier than the group treated with Antitubercular drugs only. |
| <sup>17</sup> Arun Gupta et al. Ancient Science of Life/ 2017 | 627 school going healthy Children between 5–12 years | 6 gm twice daily followed by a cup of milk | 06 months | Significant improvement in immunity (number of episodes, severity and duration of infection/allergies), energy levels, physical fitness, strength, stamina (7-point likert scale) and quality of life(KIDSCREEN QOL-27 questionnaire). |
| <sup>18</sup> J. L. N. Sastry et al. Indian Journal of Applied Research/ 2011 | 40 adults 16-65 years either healthy or with history of allergy, history of recurrent viral infections in throat, recurrent bacterial infection | 12 gm twice daily | 12 weeks | IgG and IgE levels decreased. |

The current study proved the safety and tolerability of *Chyawanprash* through non-occurence of any adverse drug reaction and comparison of hematological and bio-chemical parameters of the participants before and after the study. Despite ample scientific evidence against the efficacy of hydroxychloroquine (HCQ), health departments and physicians have continued to promote the use of HCQ both prophylactically and therapeutically.^7^ While promoting HCQ as a possible prophylactic solution to SARS-CoV-2 infection, the safety of the drug was demanded to be emphasized under closer scrutiny all over the world because of its potential to cause cardiac toxic effects and overall adverse outcomes, especially in persons with underlying coexisting conditions that increase the risk of severe COVID-19.^20^ As per the revised advisory on the use of HCQ as prophylaxis for COVID-19 infection published on the website of MoHFW, assessment of HCQ prophylaxis among 1323 HCWs indicated mild adverse effects such as nausea, abdominal pain, vomiting, hypoglycemia and cardio-vascular effects and the data from the Pharmacovigilance program of India indicated that there have been 214 reported instances of adverse drug reactions associated with prophylactic HCQ use, including 7 serious individual case safety reports with prolongation of QT interval on ECG in 3 cases.^21^ So, if HCQ is to be used, a clear informed choice needs to be offered to every contact, explaining the scarcity of evidence for its efficacy and its potential risks.^22^ Boulware et al. reported frequent mild side effects of HCQ.^23^ On the other hand, *Chyawanprash* is a *Rasayana* medicine being prescribed by Ayurveda physicians since thousands of years to improve general body strength and vitality. It is specially indicated in patients having chronic respiratory disorders. It can be administered in all age groups in every season by persons having normal digestive capacity as proven by certain studies among children.

The therapeutic value of *Chyawanprash* as an adjunct to anti-tubercular drug to augment their bioactivity and prevent their side effects is proved in clinical trials.^16,24^ *Chyawanprash* is an effective adaptogenic and antioxidant in normal people and cases of depression.^25^ It is proven that the combination phyto-chemicals offers better antioxidant effects than single antioxidant therapy.^26^ *Chyawanprash* helps to balance the three *doshas* viz., *Vata, Pitta*, and *Kapha* (bodily humors/bioenergies regulating the structure and biofunctions of the human body). In the Ayurvedic perspective, the specific actions of herbs in *Chyawanprash* are at the metabolic level.^27^ *Chyawanprash* generally helps in eliminating the accumulated stool and so in improving digestive capacity when taken regularly. This enhances the nourishment and building up of the tissues and maintaining immune homeostasis. *Chyawanprash* is a potent cardiotonic. It exerts antihyperlipidemic activity and alleviates metabolic impairments.^28,29^ *Amla*, which is the prime ingredient of *Chyawanprash*, shows antiatherogenic, anticoagulant, hypolipidemic, antihypertensive, antioxidant, antiplatelet, and vasodilatory effects, as well as lipid deposition inhibitory properties.^30,31^ *Chyawanprash* also nourishes the brain cells, harmonizes neuronal activities, improves memory, and enhances learning ability, storage, recall, and intellect. It relaxes the central nervous system (CNS), thereby acting as an anxiolytic and an antidepressive, and alleviates insomnia. Research has also suggested its procholenergic activity and antiamnesic potential.^32,33,34^

Considering all the above empirically proven benefits of *Chyawanprash*, the drug might have affected the digestion, metabolism and overall body strength or stamina of the participants in the present study. It also might have had reduced the anxiety and stress and improved sleep among HCWs. But as the study was focused to evaluate *Chyawanprash* as a safe and effective prophylactic intervention against COVID-19 infection, all the above benefits were not measured and compared. Moreover, the study was conducted at an Ayurvedic hospital functioning as COVID Care Centre which was the only feasible study setting then, when the pandemic had created panic among all and most general health care services were almost shut. So there are many chances that the participants were already consuming some Ayurvedic medicines like

Samnshani Vati, Giloy Juice, Tulsi Juice etc. as immunity boosters before getting enrolled in the trial and as the study dealt with prevention of an acute infectious disease, washout period could not be given before enrolment. The main limitations of our study were that it was done in a single COVID-19 care centre, which was a designated government hospital with a manageable patient load, good infrastructure and robust infection control practices. The risk of exposure here was not as high as general OPD where asymptomatic and mild cases with initial ILI like symptoms may be dealt before diagnosis or in other COVID-19 and non-COVID-19 public sector hospitals with a large patient burden, limited staff, inadequate personal protective equipment and frequent breaches in infection control practices. Secondly the participants were not screened for presence of anti-SARS-CoV-2 IgM and IgG antibodies before inclusion as the study was designed before the first seroprevalance survey in India.^35^

## Conclusion

*Chyawanprash* can be safely administered in health care workers but its efficacy as add on intervention to the standard measures for preventing the COVID-19 infection could be proved by clinical study conducted for long duration with larger sample size and higher dosage of the drug as had been effective in previous studies.

## Data Availability

The authors confirm that the data supporting the findings of this study are available within the article [and/or] its supplementary materials.

## Acknowledgement

We thank Dr. Geetha Krishnan Pillai, Technical Officer, TCIM Unit, WHO, who provided valuable insight and expertise that greatly assisted the drafting of the manuscript. We are grateful to Dabur India Limited for timely providing the trial drug during the pandemic.

## Authorship Confirmation

All authors contributed to this work and have reviewed and approved the manuscript prior to submission.

## Author Contributions

Conceived and designed the trial: BY, RS, AG. Acquisition of data: AG, AM, YP, RA. Analyzed the data: AT, RS. Interpretation of results: RS, PM. Manuscript drafting: PM, RS. Revised the manuscript critically: PM, RS, BY, BCSR, BSS. Final approval of the version to be published: B, NS, KSD. Accountable for all aspects of the work in ensuring that questions related to the accuracy or integrity of any part of the work are appropriately investigated and resolved: AG, AM, YP, RA, BY, BCSR, B, NS, KSD.

## References

1) Kelvin Kai-Wang To, Owen Tak-Yin Tsang, Cyril Chik-Yan Yip, Kwok-Hung Chan, Tak-Chiu Wu, Jacky Man-Chun Chan, al. Consistent Detection of 2019 Novel Coronavirus in Saliva. Clinical Infectious Diseases. 2020;71(15):841– 843 https://doi.org/10.1093/cid/ciaa149

2) World Health Organisation (WHO): Transmission of SARS-CoV-2: implications for infection prevention precautions https://www.who.int/publications/i/item/modes-of-transmission-of-virus-causing-covid-19-implications-for-ipc-precaution-recommendations, accessed 6th January 2021

3) WHO: Overview of the current COVID-19 situation https://www.who.int/countries/ind/, accessed 6th January 2021

4) COVID-19 Data Repository by the Center for Systems Science and Engineering (CSSE) at Johns Hopkins University https://github.com/CSSEGISandData/COVID-19, accessed 6th January 2021

5) WHO: Coronavirus disease (COVID-19) advice for the public: MythBusters https://www.who.int/emergencies/diseases/novel-coronavirus-2019/advice-for-public/myth-busters, accessed 6th January 2021

6) Nina PB, Dash AP. Hydroxychloroquine as prophylaxis or treatment for COVID-19: What does the evidence say?. Indian J Public Health 2020;64, Suppl S2:125–7

7) Chloroquine as a prophylactic agent against COVID-19? International Journal of Antimicrobial Agents 55 (2020) 105980 https://doi.org/10.1016/j.ijantimicag.2020.105980

8) Rathi S, Ish P, Kalantri A, Kalantri S. Hydroxychloroquine prophylaxis for COVID-19 contacts in India. Lancet Infect Dis. 2020; 20(10):1118–1119. doi: https://doi.org/10.1016/S1473-3099(20)30313-3

9) Balsari S, Sange M, Udwadia Z, COVID-19 care in India: the course to self-reliance, The Lancet Global Health, 8 (11) (2020), e1359–e1360, ISSN 2214-109X, https://doi.org/10.1016/S2214-109X(20)30384-3

10) FAQs on COVID-19 from AIIMS e-ICUs https://www.mohfw.gov.in/pdf/AIIMSeICUsFAQs01SEP.pdf accessed 21st January 2021

11) A. Kumar, D. Sathyapalan, A. Ramachandran, K. Subhash, L. Biswas, K.V. Beena, SARS-CoV-2 antibodies in healthcare workers in a large university hospital, Kerala, India, Clinical Microbiology and Infection, ]y2020, ISSN 1198-743X, https://doi.org/10.1016/j.cmi.2020.09.013.

12) Kishan Shob. Rising covid-19 infection among healthcare workforce continues to raise concerns in India. https://www.dailyrounds.org/blog/rising-covid-19-infection-among-healthcare-workforce-continues-to-raise-concerns-in-india/ dated 3.9.2020 [Accessed on 2.12.2020]

13) A. N. Uma, Dhananjay S. Kotasthane, “A Cytogenetic Study on the Efficacy of Chyawanprash Awaleha as an Antioxidant in Oral Premalignant Cancer”, Journal of Oral Oncology, vol. 2014, Article ID 864230, 5 pages, 2014. https://doi.org/10.1155/2014/864230

14) J.S. Yadav, S. Thakur & P. Chadha (2003) Chyawanprash Awaleha: A Genoprotective Agent for Bidi Smokers, International Journal of Human Genetics, 3:1, 33–38, DOI: https://doi.org/10.1080/09723757.2003.11885825

15) Gupta, N. and Mahapatra, S.C. (2013). Effects of an ancient nutritional supplement chyawanprash and vitamin c on antioxidant enzymes and serum immunoglobulin g levels in humans. Acta Hortic. 972, 61–66 doi: https://doi.org/10.17660/ActaHortic.2013.972.8

16) Debnath P.K., Chattopadhyay J., Mitra A., Adhikari A., Alam M.S., Bandopadhyay S.K., Hazra J. Adjunct therapy of Ayurvedic medicine with anti-tubercular drugs on the therapeutic management of pulmonary tuberculosis. J. Ayurveda Integr. Med. 2012;3:141–149. doi: 10.4103/0975-9476.100180.

17) Gupta A., Kumar S., Dole S., Deshpande S., Deshpande V., Singh S., Sasibhushan V. Evaluation of Cyavanapraśa on health and immunity relatedparameters in healthy children: A two arm, randomized, open labeled, prospective, multicenter, clinical study. Ancient Sci. Life. 2017;36:141–150. doi:10.4103/asl.ASL_8_17.

18) Sastry J. L. N, Gupta A, Brindavanam NB, Kanjilal S, Kumar S, Setia M, et al. Quantification of Immunity Status of Dabur Chyawanprash -A Review Part-2 (Clinical Studies). Indian J Applied Research. 2014; 4(3): p 205–211 https://www.worldwidejournals.com/indian-journal-of-applied-research-(IJAR)/fileview/March_2014_149275798161.pdf

19) Anonymous. Ayurvedic Pharmacopeia of India Part–II (Formulation); 1st ed.; Department of AYUSH: New Delhi, India, 2007; Volume I

20) Cohen M S. Hydroxychloroquine for the prevention of Covid-19 - Searching for Evidence. NEJM. August u2020;383(6):585–586

21) Revised advisory on the use of hydroxychloroquine as prophylaxis for SARS COVID19 infection. Available from: https://www.mohfw.gov.in/pdf/RevisedadvisoryontheuseofhydroxychloroquineasprophylaxisforSARSCOVID19infection.pdf Accessed 21st January 2021

22) Rathi, Sahaj et al. Hydroxychloroquine prophylaxis for COVID-19 contacts in India. The Lancet Infectious Diseases, 2020;20(10):1118 –1119 https://doi.org/10.1016/S1473-3099(20)30313-3

23) Boulware DR, Pullen MF, Bangdiwala AS, et al. A randomized trial of hydroxychloroquine as postexposure prophylaxis for Covid-19. N Engl J Med 2020;383:517–25.

24) Ojha J.K., Khanna N.N., Bajpay H.S., Sharma N. A clinical study on Chyawanprash as an adjuvant in the treatment of pulmonary tuberculosis. J. Res. Ind.Med. 1975;10:11–14.

25) Mehrotra S., Rawat A.K., Singh S. Standardization of popular ayurvedic adaptogenic preparation “Chyawanprash” and ethnokotary of its ingredients. Ethnobotany. 1995;7:1– 15

26) Liu R.H. Potential synergy of phytochemicals in cancer prevention: Mechanism of action. J. Nutr. 2004;134:3479–3485. doi: 10.1093/jn/134.12.3479S.

27) Datta Goutam K., Debnath P.K. Stress Adaptation in Ayurveda by Immunomodulatory Rasayana; Proceedings of the National Seminar on Rasayana,CCRAS; New Delhi, India. 8–10 March 1999; pp. 60–75.

28) Manjunatha S., Jaryal A.K., Bijlani R.L., Sachdeva U., Gupta S.K. Effect of Chyawanprash and vitamin C on glucose tolerance and lipoprotein profile. Ind.J. Physiol. Pharmacol. 2001;45:71–79.

29) Thakur C.P., Thakur B., Sinha P.K., Sinha S.K. The Ayurvedic medicines Haritaki, Amla and Bahira reduce cholesterol induced atherosclerosis in rabbits.Int. J. Cardiol. 1988;21:167. doi: 10.1016/0167-5273(88)90219-7.

30) Hashem-Dabaghian F., Ziaee M., Ghaffari S., Nabati F., Kianbakht S. A systematic review on the cardiovascular pharmacology of Emblica officinalisGaertn. J. Cardiovasc. Thorac. Res. 2018;10:118–128. doi: 10.15171/jcvtr.2018.20.

31) Sharma R, Martins N, Kuca K, et al. Chyawanprash: A Traditional Indian Bioactive Health Supplement. Biomolecules. 2019;9(5):161. Published 2019 Apr 26. doi:10.3390/biom9050161

32) Bansal N., Parle M. Beneficial effect of chyawanprash on cognitive function in aged mice. Pharm. Biol. 2011;49:2–8. doi: 10.3109/13880209.2010.489904

33) Sailesh K.S., Archana R., Mishra S., Symphoria Mukkadan J.K. Chyawanprash on cognitive, autonomic, and repiratory parameters in college students. Int.J. Res. Ayurveda Pharm. 2014;5:435–438.

34) Parle M., Bansal N. Antiamnesic Activity of an Ayurvedic Formulation Chyawanprash in Mice. Evid. Based Complement. Altern. Med. 2011;2011 doi:10.1093/ecam/neq021

35) SARS-CoV-2 antibody seroprevalence in India, August–September, 2020: findings from the second nationwide household serosurvey Murhekar, Manoj VAndhalkar, Rushikesh et al. The Lancet Global Health, Volume 9, Issue 3, e257–e266

